# Experiences of family caregivers regarding the health of children with congenital craniofacial anomalies in Colombia

**DOI:** 10.64898/2026.04.17.26351082

**Authors:** María Mercedes Lafaurie, Lina María Vargas-Escobar, María Clara González, Herney Alonso Rengifo

## Abstract

Recognizing the challenges faced by primary caregivers regarding the health of children with congenital craniofacial anomalies (CCAs) contributes to strengthening healthcare programs according to patientś and families’ differential needs. This qualitative study presents the experiences of 25 caregivers of children with CCAs from Bogotá and Cali, Colombia, identified from care registries and consultation statistics provideed from public high-complexity healthcare institutions. Grounded in Giorgi’s descriptive phenomenology and employing thematic analysis, this research utilized semi-structured interviews and focus groups to explore the diagnostic process and its impact, experiences with healthcare services, and the caregiver’s role and daily care activities. Data were analyzed using MAXQDA® qualitative software. Findings highlighted the emotional complexity of caring for childreńs health. Challenges included late diagnoses, pessimistic views of the children with CCAs condition by healthcare team members; lack of effective support, information, and guidance from health staff; absence of clear care and referral protocols, and limited access to specific adaptations and timely specialized care for children with CCAs. There were also reduced therapeutic services, and a pronounced gendered caregiving burden when responsibilities fall almost exclusively on mothers. System fragmentation, reflected in deficiencies in communication and a lack of clear, coordinated, and timely pathways of care, as well as the absence of adequate psychosocial support for families, emerged as common structural problems in healthcare services in both geographic settings where this research has been conducted. Gender-sensitive strategies focused on alleviating emotional concerns and the burden of caregiving from diagnosis onward within a patient and family-centered care model are decisive. Improving comprehensive CCAs training for healthcare personnel and making adjustments to care pathways are suggested to contribute to the implementation of inclusive health programs that address the diverse needs of children and their families.

## INTRODUCTION

Congenital anomalies are structural or functional abnormalities that arise during intrauterine life and can be identified prenatally, at birth, or sometimes, only later in childhood. These anomalies can be treated with medical or surgical interventions. Access to such care varies across countries and health system organizations. In childhood, congenital anomalies can result in chronic disabilities that impact children’s lives and those of their families, as well as healthcare systems and society [1].

Globally, congenital anomalies affect 8,000,000 newborns and account for 300,000 deaths annually. Its prevalence in low- and middle-income countries is 64.2 and 55.7 per 1,000 live births, respectively (i.e., 642 and 557 per 10,000 live births, respectively). This variation can be explained by social, racial, ecological, and economic factor [2]. According to a report by the National Institute of Health (INS) of Colombia, the estimated prevalence of congenital malformations was 177 cases per 10,000 live births by 2023, with a rate of 331.4 in Bogotá and 236.2 per 10,000 live births in Cali [3].

Congenital craniofacial anomalies (CCAs) are among the most prevalent pediatric conditions, and comprise disorders that affect the function of the head, face, and neck. Prevalence rates and phenotypic characteristics vary widely, and typically involve the skull, the stomatognathic system, the eyes, and/or the ears [4]. There are two major groups that CCAs can be categorized into: those caused by premature closure of the craniofacial sutures—craniosynostosis and faciocraniosynostosis—and those currently considered neurocristopathies, such as first- and second-branchial arch syndromes and orofacial clefts, including cleft lip and palate [5].

The CCAs diagnosis can have a significant impact on the psychological well-being of families [6–9]. Healthcare teams must consider the importance of the family environment surrounding the children with CCAs, adopting a comprehensive approach [10]. Additionally, healthcare professionals should also be aware of the differential needs that shape caregivers’ experiences and expectations regarding treatment [11].

A family caregiver of a child with a chronic health issue is responsible for the child’s care, performing or supervising activities of daily living, and seeking to compensate for existing dysfunctions [12]. Care involves understanding and attending to those who are unable to meet all or part of their physical, emotional, and/or affective needs [13]. Recent studies indicate that mothers are the primary caregivers of children and adolescents with chronic illness and undertake complex activities outside their usual domestic routines that stem from caring for their children. This can lead to acute and chronic physical and emotional disorders [14].

In 2023, 95.9% of Colombians were affiliated with the General Social Security Health System (SGSSS). Among them, 44.4% belonged to the contributory regime (formal employment sector), while 55.6% were in the state-supported subsidized regime. Beyond daily caregiving, many experiences of caregivers of Colombian children with CCAs are tied to care provided in public health institutions or through Health Benefit Plan Administrators (EAPB) that are state-governed [15].

This qualitative phenomenological study explored the experiences of family caregivers regarding the health of children with CCAs in Bogotá and Cali, Colombia, centering its interest on the main challenges faced by them, aiming to contribute to strengthening healthcare programs, according to patients’ and families’ differential needs.

## MATERIALS AND METHODS

### Design

This was a qualitative study using semi-structured interviews and focus groups. It was based on the descriptive phenomenology proposed by Giorgi, which focuses on experiences to discover the meaning given to everyday life and surrounding events [16]. From this epistemological perspective, the phenomenological approach is oriented toward the understanding and description of human experience in the context of people’s world. Lived experiences are understood as manifestations of the meanings and values expressed by specific individuals. Interpretation in this context is the exercise of making sense of an object of study [17].

### Context

Participants were selected from registries of pediatric patients with CCAs provided by public healthcare institutions providing high-complexity care (307 registries from Bogotá and 369 from Cali). Cities of Bogotá and Cali bring together people of different ethnicities (mainly mestizo, Indigenous, and Afro-Colombian individuals). Participants came from settings with high levels of labour informality, income instability, and a marked overburden on women within the family.

### Participants

A purposive sampling was implemented in the study. Data collection was carried out until sample saturation, which occurred when no new elements appeared in consecutive interviews and focus groups [18]. The inclusion criterion was being family caregivers of children with CCAs attending public entities of the SGSSS in the two selected cities, Bogota and Cali, in the last five years. People with limitations in verbal communication or with sensory alterations that prevented face-to-face communication were excluded. To invite them to participate, participants were contacted by telephone. Eleven of the contacted individuals expressed their decision not to participate because they felt uncomfortable sharing information about their experiences.

Overall, this study included 25 participants; 24 women (mothers) and 1 man (father). Thirteen participants were from Bogotá and 12 from Cali. The ages of their children ranged from 0 to 8 years, with the largest group being between 3 and 5 years. In regards to gender, 16 were girls and 9 were boys, contrary to what is reported in the literature where the prevalence of CCAs is higher in boys [19–20]. Ten different types of congenital craniofacial anomalies (CCAs) were reported by caregivers, with cleft lip and/or palate being the most common among the children (Table 1).

**Table 1.** Participants based on city, research technique, childreńs diagnosis, childreńs gender, and children’s age.

| Description | Criteria | Number of participants |
| --- | --- | --- |
| Participants by city | Bogotá | 13 |
|  | Cali | 12 |
| Participants by gender | Female | 24 |
|  | Male | 1 |
| Participants by research technique | Semi-structured interview | 10 |
|  | Focus group | 15 |
| Participants by children's diagnosis | Cleft lip and/or palate | 9 |
|  | Craniosynostosis | 5 |
|  | Microtia | 2 |
|  | Hydrocephalus | 3 |
|  | Microcephaly | 1 |
|  | Ectrodactyly-ectodermal dysplasia-clefting syndrome (EEC syndrome) | 1 |
|  | Pierre Robin sequence | 1 |
|  | Saethe Chozen syndrome | 1 |
|  | Apert syndrome | 1 |
|  | Muenke's syndrome | 1 |
| Participants by children's gender | Female | 16 |
|  | Male | 9 |
| Participants by children's age | 0–2 | 4 |
|  | 3–5 | 19 |
|  | 6–8 | 2 |

Source: the study

### Research Techniques

The study used semi-structured interviews and focus groups. Combining techniques enhances the credibility of the findings and offers a more comprehensive understanding of complex phenomena thought triangulation [21]. The semi-structured interviews aimed to explore the central themes of the phenomenon of interest. This technique enables researchers to uncover thoughts, feelings, and beliefs [22]. The focus groups permit us to explore aspects for which there is insufficient information and to understand how certain situations affect people [23]. The scripts guiding the interviews and focus groups addressed three general themes: the diagnosis and its impact on the family, experiences with the medical team and the health system, and caregivers’ role and daily caring activities. For each dimension the script has an open question and the necessary items or ideas to explore in the conversation. For both techniques, a pilot test was carried out.

In both cities, the interviews (average 60 minutes) were carried out privately in participants’ homes, and they could select the meeting point. The face-to-face focus group held in Bogotá (average 70 minutes) took place in a meeting room of the university leading the study. It was decided to conduct the second focus group in Bogotá and the focus group in Cali online to reduce barriers, given that caregivers faced limitations in reconciling household responsibilities with participation in the study. The online focus groups (average 90 minutes) were conducted using the Google Meet® platform. Face-to-face data collection was conducted by a female nursing professional certified in qualitative research. Online focus groups were conducted by two of the authors, both female and professors in qualitative methodology: one with a master’s degree in equality and gender (Bogota focus group), and one with a PhD in nursing (Cali focus group). Not prior relationship existed between researchers and participants. Participants signed written informed consent for their participation in the study.

### Data Analysis

In the thematic analysis based on descriptive phenomenology considered in this study, interpretation proceeds from the original data to the identification of meanings, themes and recurrent patterns. This process, as described by Sundler et al., is summarized in three steps: 1. Achieve familiarity with the data through open-minded reading; 2. Search for meanings and themes, and 3. Organizing themes into a meaningful wholeness [24]. The information collected was audio recorded, transcribed, and subsequently systematized using MAXQDA® qualitative analysis software. Three coders participated.

To guarantee the methodological rigor of the study, the following criteria were met: transferability, given the broad data collection across two geographical settings and different diagnoses as well as providing details about participants, context and methods; credibility, as the analysis was conducted with the support of MAXQDA^®^ and included a process of reflection and agreement among researchers; reliability, by characterizing the sample and presenting the data collection and analysis processes; and confirmability, verifying findings by combining two research techniques, contrasting the findings with other studies, and identifying the scope and limitations of the study [24].

### Strengths and limitations of the study

Although results can support thoughtful transfer to other settings, as Korstjens and Moser [25] point out, they do not support universal generalization, which is a limitation of the study. As for the study’s scope, it addressed 25 cases across two different geographical settings and different cultural contexts, exploring the experiences of family caregivers in caring for childreńs health with CCAs, including 10 different anomalies. Combining two research techniques is another strength of the study, which enhances data quality.

### Ethical Aspects

The research adhered to the fundamental principles established in the Declaration of Helsinki, ensuring respect for participants’ rights [26]. Additionally, all participants signed written informed consent for their participation in the study. The research protocol (PCI-2023-015) was approved by the Institutional Ethics Committee of Universidad El Bosque with Official Record # 002-2023 delivered on January 24, 2023 .

## RESULTS

As described above in the Participants section, this study included 25 participants: 24 women (mothers) and 1 man (father), 13 from Bogotá and 12 from Cali. Allowing a deeper exploration and understanding of the challenges experienced by caregivers in the context of caring for the health of children with CCAs, the information presented was organized, through a descriptive thematic analysis, into emergent themes, corresponding to the three general categories that guided the study: Diagnosis and its impact on the family, Experiences with healthcare, and Caregivers’ role and daily care activities.

### Diagnosis and its impact on the family

Recognition of the medical diagnosis of each child’s CCA was deeply mediated by emotions, family dynamics, communication with healthcare personnel, and the representations from which caregivers interpret the condition. According to reports, in both cities, the diagnostic process was marked by tensions related to the quality, timeliness, and clarity of medical information as well as the lack of guidance, empathy, and support from healthcare personnel, which generated uncertainty and sometimes complicated the coping process. These are the emergent themes: a) Late diagnoses, b) Failures in communication and information delivery, c) Pessimistic vision of the diagnosis, d) Lack of counseling and psychological support, e) Difficulties in newborn management/care, f) Depression and feelings of guilt, g) Search for information as coping strategie, h) “Holding on to God’s hand”, and i) The family as a fundamental support network.

#### a) Late diagnoses

Experiences surrounding childbirth were worsened by lack of prenatal diagnosis. The perception of late and sometimes ambiguous diagnoses generated distrust toward medical staff and had a strong emotional impact on families, as one participant noted:

*“I had two ultrasounds, and the doctors said the baby had a big head and was skinny, with low weight. Two months after birth, at the check-up, the pediatrician, who has always seen my older son, was an angel—very calm—and told me, “The baby’s head measures differently. Had anyone identified this?” I said no; I had only noticed a prominent forehead, but I saw no defects in my son—I saw him as perfect —. She ordered a test, and that was the first time I encountered the word “craniosynostosis.” I was devastated” (Mother of a child with craniosynostosis, Bogotá)*.

#### b) Failures in communication and information delivery

Significant tensions related to the quality, timeliness, and clarity of medical information were evident. Shortcomings in the provision of information emerged as critical barriers to understanding and accepting the diagnosis. As one respondent explained, medical information was unclear for her:

*“The diagnosis was explained to me. I was at a loss—I am not a doctor; I am not a nurse; I do not know medicine. He talked about mortality rates—many things I did not understand “(Mother of a child with craniosynostosis, Bogotá)*.

#### c) Pessimistic view of the diagnosis

In both geographic settings, a recurrent pattern was the medical team’s transmission of a disability-centered view of CCAs, with pessimistic prognoses and life-threatening risks, leading families to experience high levels of distress, uncertainty, and guilt. This compromises the emotional stability of caregivers and makes it difficult to build an empowered understanding of the child’s condition, as one of the participants explained:

*“In other words, there were many rejections against the child, against oneself, the father of the family, you know what I mean? In other words, every doctor said, “No, the girl will never walk” (Mother of a girl with microcephaly, Cali)*.

#### d) Lack of counseling and psychological support

Lack of counseling and psychological support negatively affected the development of healthy coping strategies, as described in some cases:

*“I think we should have counseling for each disease—each malformation as well. For example, each clinic should have a help center for certain people, because it is very difficult to be a first-time mother and have to experience something so hard” (Mother of a girl with cleft lip and palate, Cali)*.

#### e) Difficulties in basic newborn management/care

Associated with the lack of guidance, caregivers described situations denoting difficulties in the basic management/care of their newborns, with breastfeeding occupying a central place. This account reveals that situation:

*“There came a time when my son cried, and I did not know whether to hold him or not. I was afraid to breastfeed him; I was afraid something serious would happen, that he would not hear me” (Mother of a child with microtia, Bogotá)*.

#### f) Depression and feelings of guilt

Accounts associated with depression and feelings of guilt emerged strongly, adding an additional level of complexity and suffering to the caregiving process. When not addressed by healthcare personnel, these experiences interfere with initial bonding with the child and hinder the acquisition of knowledge about the child’s health condition. This is what a participant related:

*“I thought, “it was my fault.” I thought, “I didn’t take care of myself, I didn’t take the medicine.” Well, I didn’t take the vitamins. It was a clash of worlds and I thought, “It was my fault, it was my fault.” I felt that I went into a postpartum depression” (Mother of a child with microtia, Bogotá)*.

#### g) Search for information as coping strategie

One coping strategy considered effective by caregivers in dealing with the child’s diagnosis and health condition was the active search for alternative information through various means. These two accounts show us this situation:

*“I kept searching and searching until I found a pediatrician who put me in contact with the specialist” (Mother of a child with craniosynostosis and plagiocephaly, Cali)*.

*“So—pardon my words—I did not just accept it blindly. I went all in to research and be as persistent as I could” (Mother of a child with microtia, Bogotá)*.

#### h) “Holding on to God’s hand”

Families’ reactions to the diagnosis also involve the spiritual dimension as a mechanism of resilience and emotional containment, as shown in this testimony:

*“I cried—honestly, at that hospital I cried my eyes out. But in these cases, I say that family unity is something very important, and above all, holding on to God’s hand” (Mother of a child with communicating hydrocephalus, Cali)*.

##### i) The family as a fundamental support network

Regarding the role of support networks, family spaces were identified as resources for symbolic and emotional support in Cali and Bogota, as this participant noted:

*“They have been here for her since she was born—the love of the family. A little girl arrived to bring love and to unite us” (Mother of a girl with microcephaly, Cali)*.

### Experiences with healthcare

Care for children with CCAs often involves highly complex services and lengthy intervention processes that require strength and persistence from caregivers, given the barriers that can arise. Although some families were able to establish meaningful links with healthcare entities and reported that, once barriers were overcome, children underwent surgical interventions and treatments that improved their condition, the accounts also reported delays, lack of continuity, and poor coordination among key stakeholders. Emergent themes are these: a) Lack of clear pathways of care for children with CCAs, b) Difficult accessibility and timeliness of specialized care, c) Restricted access to therapeutic services, d) Limited care during the COVID-19 pandemic, e) Facilitating role of non-profit organizations, and f) Angels in white coats.

#### a) Lack of clear pathways of care for children with CCAs

The lack of clear pathways of care was revealed in several testimonies. Lack of experience in managing CCA cases was also mentioned. This not only affects the child’s health but also generates a high level of frustration, emotional exhaustion, and a sense of abandonment among caregivers. This account from a participant illustrate the situation:

*“EAPBs -*providers*-do not have a clear path of how to treat a child with this type of condition. For example, I was sent to three surgeons and, as soon as I went in, the surgeon would say “I don’t treat those cases” and wouldn’t even look at him. And the appointment was over—and I had already waited a month and a half to get it—only to request another surgeon who told me the same thing, and it happened three times. The last surgeon said, “I am training to manage those cases, but the one who really treats them is Dr. [doctor name]. So I made an appointment with him, and he was the one who performed the surgery. Even if EAPBs lack experience in certain cases, they should at least have a pathway or rely on entities that do treat such cases” (Father of a child with EEC syndrome, Bogotá).”*

#### b) Difficult accessibility and timeliness of specialized care

Caregivers described feeling exhausted when searching for specialized care for their children. Problems with timeliness were observed along with reduced access to medical appointments, as one of the respondents explains:

*Sometimes there is a lot of medical negligence because, for example, my child had to be operated on before she was six months old and the surgery was performed practically at 6 months because they did not authorize it quickly” (Mother of a girl with Muenke’s Syndrome, Cali)*.

#### c) Restricted access to therapeutic services

Access to therapies aimed at improving children’s condition was one of the critical issues mentioned in both cities. Caregivers reported difficulties obtaining authorization for therapies and a lack of continuity in therapeutic processes, as one participant noted:

*“My problem today is with therapies: they assign me therapy at an institute, I call the institute, and they say, “We’ll call you in 15 days to schedule the first appointment.” The referral expires, and that is how it has been for a while” (Mother of a girl with cleft lip and palate, Bogotá)*.

#### d) Limited care during the COVID-19 pandemic

During the COVID-19 pandemic in both cities, health restrictions led to cancelled appointments, inability to access regular medical check-ups, and implementation of virtual or home-based models that did not always adequately address families’ specific needs, generating concerns about children’s developmental progress. This account from one respondent describes what’s happened in her case:

*“It was complex because there were many restrictions. You would arrive and request an appointment and hear, “No, there’s no availability.” Sometimes the EAPBs would say you cannot enter with a companion or without a mask. These were precautions society needed, of course, but many doors were closed—many programs and many access points to healthcare—so I think it was complex, very complex, on the medical side*” (Mother of a girl with microtia, Bogotá).

#### e) Facilitating role of non-profit organizations

Several cases highlighted the importance of non-profit organizations that provided surgical procedures, medications, or emotional support at no cost, making a significant difference in cases that public entities were unable to solve in a timely manner, as this account reveals.

*“Well, the institution that has welcomed him is XXX (name of the institution). That is the only one I was told of, and I started to inquire. Then they accepted me” (Mother of a child with hydrocephalus, Cali)*.

#### f) Angels in white coats

Given the multiple hurdles caregivers must overcome to secure care for their children, finding a competent and committed professional is especially valued and sometimes viewed as encountering someone with a special spiritual power who appears to change the situation. These two participants describe their experiences:

*“That same day he was born, Dr. XXX (name of the doctor) visited him in the ICU and diagnosed him. That same day, his diagnosis was confirmed. Fortunately, Dr. XXX was there; he gave me all the orders for surgery. They performed mandibular distraction, and on his first day of life, he underwent surgery. ” (Mother of a child with Pierre Robin Sequence, Cali)*.

*“Thank God I arrived at that Hospital; I love Dr. XXX (name of the doctor). He has been a light in my life and a guide. I mean, my daughter’s surgery—today you can’t even tell she had a condition (Mother of a girl with cleft lip and/or palate, Bogotá)*.

### Caregivers’ role and daily care activities

The stories from Bogotá and Cali show that the daily care of children with CCAs is a complex and demanding experience that impacts caregivers’ emotional, social, and work life. In both contexts, the role of the caregiver is constructed as an integral function that ranges from managing medical appointments, therapies and treatments, to permanent support in the basic activities of daily living. From this category emerged the next themes: a)Caring for children with CCAs: an absorbing task, b) Caregiving is emotionally charged, c)Gender overload, and d) Development of children with CCAs: an ongoing challenge.

#### a) Caring for children with CCAs: an absorbing task

The testimonies often emphasize the logistical and care work linked to maintaining medical and functional routines, and they highlight the absorbing and all-encompassing nature of the role, which frequently reconfigures caregivers’ personal and professional projects, as this participant notes:

*“In other words, you do not have caregiving support; you are a 100% caregiver. Yes, 100%” (Mother of girl with craniosynostosis Cali)*

#### b) Caregiving is emotionally charged

The subjective impact of caregiving becomes more visible, underscoring the emotional burden of this role and the caregiver’s physical and mental strain, as these accounts reveal:

*“The surgery, the recovery, and the time they are in the operating room—those are truly moments of great anguish” (Mother of a child with cleft lip and palate, Bogotá)*.

*“You have to keep ten eyes on him all the time, because he talks to you, he gets excited, he laughs, he runs all the time, so you’re constantly saying, “Careful there,” “Watch the edge of the wall,” “Be careful—don’t grab the ball.” But it’s more our fears than what he needs, because he is a child who does not require much care—other than a protective helmet when he goes to the park—because his skull still hasn’t closed” (Mother of a child with craniosynostosis, Bogotá)*.

#### c) Gender overload

In both cities, caregivers assume a sustained responsibility, which is rarely shared or alleviated by institutional or community networks. Gender overload is evident, as women mostly perform this role, juggling multiple simultaneous tasks without formal recognition or sufficient structural support. This is the view of one of the participants:

*“If I go to work or for a walk, who do I leave her with? Obviously, I’m going to be stressed—I’m going to be anxious. Why? Because I say, “No, I know my daughter is not going to be okay.” So, I prefer to work independently from home and take care of her myself” (Mother of a girl with microcephaly, Cali)*

#### d) Development of children with CCAs: an ongoing challenge

Caring for a child with CCA is a challenge for caregivers, a daily struggle that is rewarded by seeing their children progress, as this participant explained:

*“When a child is born with a condition, it is additional work, let’s put it that way, for a family, isn’t it? So yes, sometimes one feels frustrated for having studied and not being able to fulfill oneself professionally, but the comforting thing is to see them succeed, even if it is just a small piece of what they can conquer” (Mother of a child with cleft lip and/or palate, Bogotá)*.

## d) DISCUSSION

Results of this study are consistent with the literature, regarding the challenges faced by caregivers of children with CCAs, such as craniosynostosis, craniofacial microsomia, and cleft lip and/or palate, which involve the complexities of the health condition itself as well as the social and psychoemotional factors embedded in the caregiving experience. The analysis shows that understanding the children’s condition cannot be assumed as a linear process of information-transmission, but rather as a relational, situated, and affective construction.

The presence of anxiety, stress, feelings of guilt, and depressive symptoms in caregivers, which emerged through the analysis, is highlighted in previous studies [27–28]. Caregivers of children with CCAs experience psychoemotional effects, especially in the initial stages of diagnosis, which underscores the need to address their mental health from the time of detection as part of a comprehensive approach [27–28]. The late diagnosis reported by different participants, which can be associated with limitations in implementing the maternal–perinatal care pathway, particularly affected caregivers’ experiences. Early detection not only supports medical decision-making but also reduces the impact of having a child with CCAs [29]. Communication about the diagnosis by healthcare providers is a critical issue, according to participants in this study, which is discussed by authors such as Stock et al. [28] and Johns et al. [30].

Another common finding shared with other research is the lack of emotional support for caregivers of children with CCAs from interdisciplinary teams [28, 30]. Moreover, the information and guidance received from healthcare professionals is often perceived by participants as unclear or of insufficient quality, as is remarked by some authors [30–31]. Consequently, caregivers in this study turned to coping strategies such as actively seeking information on their own. It is noted that when these strategies occur without a structured network of professional support, greater affective involvement may be generated [27, 32]. In this sense, it is essential that healthcare networks include timely, high-quality, accessible, and differential care for children with CCAs and their families, where understanding the psychoemotional experiences of patients and caregivers is central [33–34].

Difficulties accessing specialized programs, as detected in our research, are also reported by Sawasdipanich et al. [31] in Thailand. As has been seen by authors, caregivers of children with CCAs must navigate complex and bureaucratic systems that hinder treatments. On the other hand, Johns et al. [30] revealed challenges in accessing intervention programs and locating experienced surgeons, as well as unmet care coordination needs in some of the experiences within a national sample of U.S. caregivers of children with microsomia. Our findings suggest an ambivalent and fragmented experience regarding access to institutional support for the care of children with CCAs, whose complex health conditions require effective and timely referral pathways involving interdisciplinary teams and highly specialized consultation. These resources are restricted within institutions from Colombian health system, where poor coordination in service delivery and fragmentation of care are often observed [35]. Developing adjustments in care pathways and consolidating guidelines addressing patients’ and familieś differential needs is imperative.

Caregivers assume an active role in monitoring their children’s health, which involves managing administrative procedures, coordinating medical appointments, supervising compliance with treatments and administering medications. These actions are commonly aimed at achieving timely and continuous access to diagnostics, medical and therapeutic services. Their challenges may be alleviated by healthcare teams, reducing burden of care, using warm family-centered supporting style, promoting coping and linking caregivers to educative and support services [29]. In both settings, Bogota and Cali, most of the caring responsibilities fall almost exclusively on mothers, who not only provide direct care for their children but also carry all or most household tasks. This double burden is closely linked to traditional gender norms that continue to position women as natural caregivers and household managers—often without support or equitable redistribution—a finding consistent with other authors like Macedo et al. [14] and Johns et al. [29]. This caregiving experience, far from being limited to a set of functional tasks, is deeply intertwined with women’s identity, emotions, and life choices.

Multidisciplinary teams can play a significant role in supporting positive adjustment in patients affected by craniofacial conditions and their families from birth [29–30]. Considering the findings of the study, the training of healthcare professionals—beginning in academia—should include specialized clinical communication, and psychosocial skills for the humanized management of children with CCAs to provide person and family-centered care that recognizes and supports the emotional, informational, and practical challenges faced by caregivers of children with these conditions. This applies across all levels of complexity. Specialist and generalist healthcare professionals require adequate academic preparation and training for manage CCAs in children, particularly in terms of guidance, communication, education, and accompaniment throughout the care process, implementing gender-sensitive and patient and family-centered multidisciplinary approaches. It contributes to the development of inclusive health programs that address the differential needs of children with CCAs and their families, including early diagnosis.

## e) CONCLUSIONS

Delayed diagnoses, limited high-quality communication and guidance, and system fragmentation, reflected in restricted access to specific adaptations and specialized care for children with CCAs, as well as the absence of adequate psychosocial support for families, emerged as common structural problems in this study when gendered caregiving burden is evident. Therefore, strategies are required to enable caregivers—and subsequently, the children and adolescents themselves—to navigate structured, continuous, flexible, and timely care pathways tailored to their specific needs. Psychosocial and gender-sensitive strategies focused on alleviating the emotional burden of caregiving from diagnosis onward within a patient and family-centered care model are decisive. Improving comprehensive CCAs training for healthcare personnel and consolidating guidelines addressing patients’ and their familieś differential needs are suggested to contribute to the implementation of inclusive health programs in public-provided high-complexity care entities in both geographic settings where this research has been implemented.

Exploring the lived experiences of children and adolescents with CCAs by delving deeper into their support needs in healthcare, as well as identifying successful family-centered care interventions for children with craniofacial conditions and their caregivers, could help fill a research and practice gap.

## Data Availability

Data available on request.

## ACKNOWLEDGEMENTS

The authors thank the participants for their contributions to interviews and focus groups and public institutions Hospital Simon Bolivar-Subred Norte E.S.E., Universidad del Valle and Hospital Universitario del Valle for their collaboration with the project.

## REFERENCES

1. World Health Organization. Congenital disorders. Geneva, Swiss: WHO; 2023.

2. Al-Dewik N, Samara M, Younes S, Al-Jurf R, Nasrallah G, Al-Obaidly S, et al. Prevalence, predictors, and outcomes of major congenital anomalies: A population-based register study. Sci Rep. 2023; 13(1):2198. 10.1038/s41598-023-27935-3

3. Instituto Nacional de Salud. Informe de evento defectos congénitos Colombia (Birth Defect Event Report Colombia). Bogota, Colombia: INS; 2023.

4. Myhre A, Agai M, Dundas I, Billaud Feragen K. All eyes on me: A qualitative study of parent and patient experiences of multidisciplinary care in craniofacial conditions. Cleft Palate Craniofac J. 2019; 56(9):1187–1194. 10.1177/1055665619842730

5. Sorolla JP. Anomalías craneofaciales. Revista Médica Clínica Las Condes. 2009. 21(1): 5–15. 10.1016/S0716-8640(10)70500-9

6. Santos AJ, Braz P, Folha T, Machado A, Matías-Dias C. Parents of children diagnosed with congenital anomalies or cerebral palsy: Identifying needs in interaction with healthcare services. Children (Basel). 2023; 10(6):1051. 10.3390/children10061051

7. Marcus E, Latos-Bielenska A, Jamry-Dziurla A, Barišić I, Cavero-Carbonell C, Den Hond E, et al. Information needs of parents of children with congenital anomalies across Europe: A EUROlinkCAT survey. BMC Pediatr. 2022; 22:657. 10.1186/s12887-022-03734-z

8. Shapiro DN, Waljee J, Buchman S, Ranganathan K, Warshcausky S. Gender views and relationships in families of children with craniofacial differences. Cleft Palate Craniofac J. 2018; 55(2):189–195. 10.1177/1055665617726534

9. Stock NM, Costa B. Provision of care for families affected by craniofacial conditions: The views of nonspecialist health professionals. Cleft Palate Craniofac J. 2019; 57(4):470–476. 10.1177/1055665619883151

10. Myhre A, Råbu M, Feragen KB. Are we together in this? Relationship experiences of parents of children with craniofacial anomalies. Cleft Palate Craniofac J. 2024; 61(10):1646–1656. 10.1177/10556656231180512

11. Johns AL, Luquetti DV, Brajcich M, Heike C, Stock N. In their own words: caregiver and patient perspectives on stressors, resources, and recommendations in craniofacial microsomia care. J Craniofac Surg. 2018; 29(8): 2198–2205. 10.1097/SCS.0000000000004867.

12. Barrera L, Blanco L, Figueroa P, Pinto N, Sánchez B. Habilidad de cuidadores familiares de personas con enfermedad crónica. Mirada internacional. Aquichan. 2006; 6(1):22–33.

13. Inter-American Commission of Women. COVID-19 in Women’s Lives: The Global Care Emergency. Washington, D.C., USA: Organization of American States; 2020.

14. Macedo EC, da Silva LR, Paiva MS, Pereira MN. Burden and quality of life of mothers of children and adolescents with chronic illnesses: an integrative review. Rev Lat Am Enfermagem. 2015; 23 (4):769–77. 10.1590/0104-

15. Republic of Colombia. National Department of Statistics. National Quality of Life Survey – ECV; 2023.

16. Martínez-Ávila B, Álvarez-Aguirre A. Application of the phenomenology of Amedeo Giorgi as methodological support. ACC CIETNA. 2021; 8(1):106–12. doi:10.35383/cietna.v8i1.570

17. Giorgi A, Giorgi B, Morley J. The Descriptive Phenomenological Psychological Method. In: Willig C, Rogers WS (Eds.). The Sage Handbook of Qualitative Research in Psychology. Edition: 2nd, Chapter: 11. New York, USA: Sage Editors; 2017. 10.4135/9781526405555.n11

18. Ahmed SK. Sample size for saturation in qualitative research: Debates, definitions, and strategies. JMSPH. 2025; 5: 100171. 10.1016/j.glmedi.2024.100171

19. Jahanbin A, Jamalinasab A, Niazi AE. Variations in orofacial clefts. J Craniofac Surg. 2021; 32(2): e179–e182. 10.1097/SCS.0000000000007027.

20. Silva-Giraldo X, Porras-Hurtado GL. Characterization of congenital craniofacial anomalies in a specialized hospital of Risaralda, Colombia. 2010-2014. Rev.Fac.Med. 2018; 66 (2): 223–227. 10.15446/revfacmed.v66n2.61551.

21. Flick U. The SAGE Handbook of Qualitative Research Design, Vol 2. New York, USA: Sage Editors; 2022 10.4135/9781529770278

22. DeJonckheere M, Vaughn LM. Semistructured interviewing in primary care research: a balance of relationship and rigor. Fam Med Community Health. 2019;7 (2): e000057. 10.1136/fmch-2018-000057

23. Hamui-Sutton A, Varela-Ruiz M. La técnica de grupos focales. RIEM. 2013; 2(5):55–60.

24. Sundler AJ, Lindberg E, Nilsson C, Palmér. Qualitative thematic analysis based on descriptive phenomenology. Nurs Open. 2019; 6(3), 733–739. 10.1002/nop2.275.

25. Korstjens I, Moser A. Series: Practical guidance to qualitative research. Part 4: Trustworthiness and publishing. The European journal of general practice. 2018; 24(1), 120–124. 10.1080/13814788.2017.1375092

26. World Medical Association. WMA Declaration of Helsinki – Ethical Principles for Medical Research Involving Human Participants. Ferney-Voltaire, France: WMA; 2024.

27. Costa B, Edwards W, Wilkinson-Bell K, Stock NM. Raising a child with craniosynostosis: Psychosocial adjustment in caregivers. Cleft Palate Craniofac J. 2023; 60 (10):1284–1297. 10.1177/10556656221102043.

28. Stock NM, Costa B, Parnell J, Johns A, Crerand C, Feragen KB, et al. A conceptual thematic framework of psychological adjustment in caregivers of children with craniofacial microsomia. Cleft Palate Craniofac J. 2024; 62 (7): 1–13. 10.1177/10556656241245284.

29. Chen J, Kanekar S. Imaging of congenital craniofacial anomalies and syndromes. Clin Perinatol. 2022;49(3):771–790. 10.1016/j.clp.2022.04.005

30. Johns AL, Stock NM, McWilliams D, Rahman M, Costa B, Crerand CE, et al. Embarking on a Treatment Journey: Experiences of Caregivers of Young Children with Craniofacial Microsomia. J Craniofac Surg. 2025; 36(7). 10.1097/SCS.0000000000011643..

31. Sawasdipanich N, Chaithat B, Dangsomboon A, Rojvachiranonda N. Perceptions of caregivers regarding healthcare service accessibility of children with craniofacial anomalies: A qualitative descriptive study. Pac. Rim Int. J. Nurs. Res 2025; 29 (2):231–245. 10.60099/prijnr.2025.271221.

32. Feragen KB, Stock NM, Myhre A, Due-Tønnessen BJ. Medical stress reactions and personal growth in parents of children with a rare craniofacial condition. Cleft Palate Craniofac J. 2020; 57(2):228–237. 10.1177/1055665619869146.

33. Feragen KB, Myhre A, Stock NM. “Exposed and vulnerable”: Parent reports of their child’s experience of multidisciplinary craniofacial consultations. Cleft Palate Craniofac J. 2019; 56 (14):1230–1238. 10.1177/1055665619851650.

34. Chi Adam KB, Hariri F, Murat NA, Yin LM, Amran MN. Parental experiences in the management of children with syndromic craniofacial diagnoses in Malaysia. IMJM Medical Journal Malaysia. 2021; 20 (3):53–59. 10.31436/imjm.v20i3

35. Chavez, B.M. Contributions to the transformation of the Colombian health system. Rev. Fac. Nac. Salud Pública. 2023; 41(1): e348269. 10.17533/udea.rfnsp.e348269

